# Factors associated with the readiness assessment of health facility services in Yaoundé, Cameroon

**DOI:** 10.64898/2026.06.30.26356973

**Authors:** Ghislaine Octavia Tedjo Pokam, Collins Buh Nkum, Daniele Sandra Yopa, Marie Nicole Ngoufack, Carole Debora Nounkeu, Gilles Protais Lekelem Dongmo, Palmer Masumbe Netongo, Georges Eric Nseme Etouckey, Georges Nguefack-Tsague

## Abstract

**Introduction:** Reliable information on service readiness is essential for strengthening health systems and advancing universal health coverage. In Cameroon, structural imbalances and the predominance of private-sector facilities raise concerns regarding the operational capacity of health facilities to deliver essential services. This study aimed to determine factors associated with health facility readiness in Yaoundé to inform evidence-based policy and service delivery improvements.

**Methods:** A cross-sectional analytical study was conducted from November 4 to December 27 2024 among health facilities in Yaoundé. Data were collected using the World Health Organization Service Availability and Readiness Assessment (SARA) tool. Readiness was measured across five domains (1) trained staff and guidelines; (2) essential equipment; (3) standard precautions for infection prevention; (4) diagnostic capacity; and (5) essential medicines), comprising 47 tracer items. Facilities scoring ≥80% were classified as having good readiness. Bivariate analyses and multivariate logistic regression were performed to identify factors associated with good readiness.

**Results:** A total of 205 health facilities were surveyed; most were urban (97.6%), private secular (89.8%), and categorized as 6th level (86.8%). Overall, 57.1% (117/205) achieved good readiness. Readiness varied significantly across health districts (p=0.015), with Efoulan (78.8%) performing highest. In multivariate analysis, *absence of Prevention of Mother-to-Child Transmission (PMTCT) services* (aOR=0.17; 95% CI: 0.05–0.55; p=0.003) and *absence of childbirth services* (aOR=0.18; 95% CI: 0.06–0.55; p=0.003) were independently associated with lower odds of good readiness.

**Conclusions:** Only slightly more than half of facilities in Yaoundé met the operational readiness benchmark. Availability of PMTCT and childbirth services appears to be a strong indicator of broader facility preparedness. Strengthening maternal and child health service capacity may serve as a strategic entry point for improving overall facility readiness and advancing equitable health system strengthening in Cameroon.

## Introduction

The management, monitoring and evaluation of health systems must be based on solid information concerning the supply and quality of health services[1]. Global health initiatives have underscored the need for reliable systems to monitor health service capacity at national levels, encompassing public and private sectors, to measure the operational readiness of facilities to deliver essential interventions[2]. Despite growing investments, many low- and middle-income countries (LMICs) lack up-to-date, accurate information on health system inputs and the extent to which facilities are prepared to provide quality services[3].

A core function of any health system is to provide access to quality care. Service readiness, the capacity of health facilities to deliver services is a prerequisite for quality of care. It requires the presence of trained staff, guidelines, infrastructure, equipment, medicines, and diagnostics[4]. Assessing readiness is therefore critical for identifying gaps and guiding resource allocation.

In Cameroon, the national health system faces significant structural challenges. The overall density of health facilities is approximately one facility per 4,227 inhabitants across all sectors, and one public facility per 9,113 inhabitants[5]. Public facilities constitute only a small proportion of total facilities nationally. In fact, the provision of health care and services in this large metropolis is dominated by private, secular, for-profit health facilities. In Yaoundé, approximately 85% of facilities are private, secular, for-profit institutions. This pattern varies substantially across regions. Public sector coverage is moderate in the South (66%), South-West (63%), North-West (60%), Center excluding Yaoundé (55%), West (54%), and Littoral excluding Douala (52%) regions. Conversely, the North and Far North regions depend heavily on public facilities, which constitute 86% of all facilities[6]. These disparities reflect underlying inequities in healthcare access with important implications for health system planning.

In response, Cameroon updated its national health map in 2021, replacing the previous decade-old version. However, significant problems persist. National standards recommend one health district per 50,000 inhabitants, yet Efoulan Health District in Yaoundé serves nearly one million residents a twenty-fold exceedance. To meet standards, this district alone requires subdivision into at least five districts, and Yaoundé would need approximately 20 health districts rather than the current eight[7].

Building on international frameworks like the Paris Declaration and IHP+, global partners have developed a comprehensive approach for monitoring and evaluating health systems strengthening[8]. This emphasizes strengthening data platforms to improve information availability and utilization for health sector decision-making[9].

The Service Availability and Readiness Assessment (SARA) methodology used in this study aligns with this framework, providing a standardized approach to data collection that can be combined with records reviews to assess data quality. Given these contextual challenges and the need for evidence-based health system planning, this study aimed to determine the factors associated with health facility service readiness in Yaoundé, Cameroon.

## Methods

### Design

This study employed an analytical cross-sectional design with three specific aims: to describe health facility characteristics, to evaluate service availability, and to measure the service readiness index. These analyses were conducted to identify factors associated with health facility readiness in Yaoundé.

### Ethical considerations

Ethical approval was obtained from the Centre Regional Ethics Committee for Human Health Research (CE N^0^ 01090/CRERSHC/2024 of October 8^th^ 2024). Additional authorization was secured from the district medical officers for each of the eight health districts. All participants provided written informed consent after receiving detailed information about the study’s purpose, procedures, risks, and benefits. Data were anonymized to ensure confidentiality.

### Study Setting

The study was conducted in the city of Yaoundé, the capital of Cameroon, located in the Centre region. Yaoundé is a metropolis with nearly 5 million inhabitants and comprises eight health districts: Biyem-Assi, Cité Verte, Djoungolo, Efoulan, Mvog-Ada, Nkolbisson, Nkoldongo, and Odza. Cameroon’s health system faces several challenges, including unbalanced and sparse health coverage, high professional workload for nursing staff, and significant constraints in accessing health structures. In some health districts, there is one doctor per 500,000 inhabitants or one nurse per 140,000 inhabitants, far below international standards recommending one doctor per 10,000 inhabitants and one nurse per 5,000 inhabitants[10].

### Study Period and participants

This study was conducted from November 4 to December 27 2024. Our study targeted all functioning health facilities within the eight health districts of Yaoundé. A sample of 205 health facilities was drawn from the master facility list (MFL), which contained 666 facilities.

### Sampling Strategy

#### Sampling frame

The sampling frame for this study was the Yaoundé Master Facility List (MFL), maintained by the Ministry of Public Health of Cameroon, which enumerated all registered health facilities across the eight health districts of Yaoundé at the time of study initiation. The MFL comprised a total of N = 666 health facilities, spanning public, private secular (for-profit), and private denominational (faith-based, non-profit) ownership types across the districts of Biyem-Assi, Cité Verte, Djoungolo, Efoulan, Mvog-Ada, Nkolbisson, Nkoldongo, and Odza. The MFL was cross-validated against district health office records and updated immediately prior to sampling to minimize coverage error.

#### Sampling Design

A stratified random sampling design was employed to ensure proportional representation of health facilities across key structural dimensions. This approach was selected over simple random sampling to guarantee adequate representation of each health district and ownership type.

The sampling frame was first stratified by health district (eight strata: Biyem-Assi, Cité Verte, Djoungolo, Efoulan, Mvog-Ada, Nkolbisson, Nkoldongo, Odza) and then by facility ownership type within each district (public; private secular; private denominational). Within each stratum, individual health facilities were selected by simple random sampling without replacement, using computer-generated random numbers. The number of facilities allocated to each stratum was proportional to the stratum’s share of the total MFL.

#### Sample Size Estimation

The sample size (n) of the healthcare facilities to consider for this study was computed using Cochran’s modified formula for finite populations[11]: 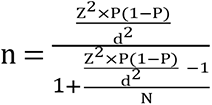 where Z is the approximate value of the 97.5 percentile point of the standard normal distribution =1.96, P is the proportion of healthcare facilities having good readiness=0.5 (default due to non-availability), d is the precision= 0.06, N=666 (total number of healthcare facilities in Yaoundé), and 5% non-response rate. The numerical application of this formula gives the minimum sample size (n) of 200 healthcare facilities to be visited.

### Inclusion criteria

All health facilities listed in the Yaoundé MFL were eligible for inclusion if they: (i) were located within the administrative boundaries of one of the eight health districts of Yaoundé; (ii) were operational (i.e., providing health services to patients) at the time of the field visit; and (iii) whose person in charge, or the most senior staff member available, provided written informed consent to participate.

### Exclusion criteria

Facilities were excluded if they: (i) had permanently closed or were temporarily suspended from service delivery at the time of the field visit; (ii) were under construction and had not yet commenced patient services; (iii) whose person in charge refused participation; or (iv) could not be located at the registered address in the MFL after two attempted visits.

### Data Collection Procedure

#### Instrument

Data were collected using a structured questionnaire adapted from the standard *Service Availability and Readiness Assessment (SARA)* tool developed by the World Health Organization[12]. For this study, the standard questionnaire was contextualized to the Cameroonian setting through adaptation of: facility category classifications consistent with national health system nomenclature; institutional management hierarchies; national clinical guidelines and service protocols; cadre-specific personnel categories; and the national essential medicines list (including disease-specific lists for HIV/AIDS and tuberculosis). These adaptations did not alter the core domains or tracer items of the SARA tool, preserving cross-study comparability.

The adapted questionnaire was pre-tested in five health facilities not included in the main sample, located in Yaoundé, to assess clarity, length, and cultural appropriateness. The final instrument comprised 47 tracer items spanning five readiness domains.

#### Data Collection Approach

Data were collected through a combination of two complementary methods, in accordance with standard SARA protocol: (i) structured interviews with the facility head or, in their absence, the most senior clinically qualified staff member available; and (ii) direct observation of tracer items (equipment, medicines, guidelines, and infection prevention supplies) on the day of assessment. This combined approach was used to minimise social desirability bias and to verify self-reported availability of key inputs.

Field data collection was conducted by trained data collectors using electronic data capture on Android tablets running Google Forms, with printed questionnaires retained as backup. Data collectors received standardised training covering SARA methodology, interviewing techniques, facility navigation procedures, and quality assurance protocols. Each completed form was reviewed on the same day by the supervising investigator for completeness and internal consistency. Material resources included tablets, memory cards, printed questionnaires, stationery, and informed consent forms. All facilities were visited in person; data collectors obtained written informed consent before administering the questionnaire

#### Measurement of Readiness Assessment

The primary outcome was overall facility service readiness, operationalized as a composite score derived from the five SARA core domains: (1) trained staff and guidelines; (2) essential equipment; (3) standard precautions for infection prevention; (4) diagnostic capacity; and (5) essential medicines. Each of the 47 tracer items was scored binarily: 1 if the item was present and functional on the day of assessment, or 0 if absent or non-functional. A domain score was calculated as the proportion of present-and-functional tracers for that domain, expressed as a percentage. The overall readiness score was the unweighted arithmetic mean of the five domain scores, also expressed as a percentage[13].

Facilities scoring ≥80% were classified as having ’good readiness’, and those scoring <80% as having ’limited readiness’. The 80% threshold was selected a priori as an operational benchmark widely applied in SARA-based assessments to indicate an acceptable level of service readiness, although no universal cut-off has been formally established by WHO.

### Statistical Analysis

#### Data management

Data collected were exported to Microsoft Excel 2013 for cleaning, verification, and coding. Cleaning involved checking for missing values, out-of-range entries, and logical inconsistencies. Tracer items were coded binarily (1 = present and functional; 0 = absent or non-functional). Domain scores and the overall readiness index were computed in Excel before the dataset was exported to IBM SPSS Statistics version 27.0 (IBM Corp., Armonk, NY, USA) for all statistical analyses.

#### Descriptive analysis

Descriptive statistics were used to characterize health facilities and survey respondents. Categorical variables (location, district, category, ownership, manager qualification, manager sex, service availability) are presented as frequencies and percentages. Continuous variables (respondent age, years of service) were assessed for normality using the Shapiro-Wilk test and are reported as mean ± standard deviation (SD) where normally distributed, or as median with interquartile range (IQR) where skewed. Readiness domain scores and the overall readiness index were presented as means with SDs and as proportions exceeding the 80% threshold.

#### Bivariate analysis

The association between each independent variable (facility characteristics and service availability indicators) and the binary primary outcome (good readiness ≥80% vs. limited readiness <80%) was assessed using Pearson’s chi-square test, or Fisher’s exact test where any expected cell count was less than 5. Crude odds ratios (OR) with 95% confidence intervals (CI) were computed. Variables with a p-value <0.20 in bivariate analysis were considered candidates for inclusion in the multivariable model, to avoid prematurely excluding potentially confounded associations at the conventional α = 0.05 threshold.

#### Multivariable analysis

Binary multiple logistic regression was performed to identify factors independently associated with good readiness, adjusting for potential confounders identified in bivariate analysis. Results were reported as adjusted odds ratios (aOR) with 95% CIs and two-sided p-values. Model calibration was assessed using the Hosmer-Lemeshow goodness-of-fit test; a p-value >0.05 was considered indicative of adequate fit. Variables were entered simultaneously (enter method). Statistical significance was set at α = 0.05 for the final model.

## Results

### Characteristics of health facilities and respondents

A total of 205 health facilities were enrolled and completed the assessment. Table 1 summaries their key characteristics. The vast majority were located in urban areas (97.6%), reflecting the predominant urban geography of Yaoundé. Private secular (for-profit) facilities constituted 89.8% of the sample, with public (4.4%) and private denominational (faith-based, 5.8%) facilities together accounting for the remainder. Sixth-category (primary care) facilities made up 86.8% of the sample, consistent with the level structure of the Yaoundé health system.

**Table 1.** Characteristics of health facilities.

| **Variable** | **Category** | **n** | **%** |
| --- | --- | --- | --- |
| Location | Urban | 200 | 97.6 |
|  | Rural | 5 | 2.4 |
| Health District | Biyem-Assi | 55 | 26.8 |
|  | Efoulan | 33 | 16.1 |
|  | Mvog-Ada | 32 | 15.6 |
|  | Nkolbisson | 23 | 11.2 |
|  | Cité Verte | 20 | 9.8 |
|  | Djoungolo | 19 | 9.3 |
|  | Nkoldongo | 17 | 8.3 |
|  | Odza | 6 | 2.9 |
| Facility Category | 6th Category | 179 | 86.8 |
|  | 5th Category | 22 | 10.7 |
|  | 4th Category | 5 | 2.4 |
| Ownership | Private secular | 184 | 89.8 |
|  | Private denominational | 12 | 5.8 |
|  | Public | 9 | 4.4 |
| Manager Qualification | State Registered Nurse | 110 | 53.7 |
|  | Medical Doctor | 45 | 22.0 |
|  | Laboratory Technician | 20 | 9.8 |
|  | Assistant Nurse | 5 | 2.4 |
|  | Other | 25 | 12.1 |
| Manager Gender | Female | 105 | 51.2 |
|  | Male | 100 | 48.8 |

The largest shares of sampled facilities were located in the Biyem-Assi (26.8%), Efoulan (16.1%), and Mvog-Ada (15.6%) health districts; the smallest share was in Odza (2.9%). Among facility managers, more than half were State Registered Nurses (53.7%), and the sex distribution was nearly balanced (female 51.2%, male 48.8%).

Among survey respondents (who were not always the same individual as the facility manager), the majority were also State Registered Nurse (65.3%), predominantly female (67.3%), with a mean age of 36 years (SD 10; range 21–76) and a mean of 9 years of professional experience (SD 7; range 1–47).

### 2. Overall readiness assessment

Applying the pre-specified 80% operational threshold, 117 of 205 facilities (57.1%) achieved good readiness (≥80%), while 88 (42.9%) demonstrated limited readiness (<80%; Table 2). This indicates that nearly two in five health facilities in Yaoundé lacked the minimum essential inputs required for adequate service delivery at the time of assessment.

**Table 2.** Overall Health Facility Readiness (n = 205)

| Readiness Status | n | % |
| --- | --- | --- |
| $\geq 80\%$ (Good readiness) | 117 | 57.1 |
| $< 80\%$ (Limited readiness) | 88 | 42.9 |

### Factors associated with the quality of HFs services

Table 3 presents the bivariate associations between facility characteristics and readiness status. Health district was the only structural variable significantly associated with good readiness (p = 0.02). Efoulan district had the highest proportion of facilities achieving good readiness (78.8%), followed by Cité Verte (70.0%) and Nkolbisson (69.6%). The lowest-performing districts were Djoungolo (31.6%) and Nkoldongo (35.3%). Facility category (p = 0.13), ownership type (p = 0.89), location (p = 0.09), and manager sex (p = 0.66) were not significantly associated with readiness in bivariate analysis.

**Table 3.** Bivariate Analysis of Facility Characteristics and Good Readiness (≥80%)

| Variable | Category | Limited<br>readiness <80%,<br>n (%) | Good<br>readiness<br>$\geq 80\%$, n (%) | p-value |
| --- | --- | --- | --- | --- |
| Location | Urban | 85 (42.5) | 115 (57.5) | 0.09 |
|  | Rural | 4 (80.0) | 1 (20.0) |  |
| Health district | Biyem-Assi | 27 (49.1) | 28 (50.9) | 0.02 |
|  | Cité Verte | 6 (30.0) | 14 (70.0) |  |
|  | Djoungolo | 13 (68.4) | 6 (31.6) |  |
|  | Efoulan | 7 (21.2) | 26 (78.8) |  |
|  | Mvog-Ada | 15 (46.9) | 17 (53.1) |  |
|  | Nkolbisson | 7 (30.4) | 16 (69.6) |  |
|  | Nkoldongo | 11 (64.7) | 6 (35.3) |  |
|  | Odza | 3 (50.0) | 3 (50.0) |  |
| Facility category | 6th | 82 (45.8) | 97 (54.2) | 0.13 |
|  | 5th | 5 (22.7) | 17 (77.3) |  |
|  | 4th | 2 (40.0) | 3 (60.0) |  |
| Ownership | Private secular | 81 (43.8) | 104 (56.2) | 0.89 |
|  | Private denominational | 4 (33.3) | 8 (66.7) |  |
|  | Public | 3 (50.0) | 3 (50.0) |  |
| Manager sex | Female | 44 (41.9) | 61 (58.1) | 0.66 |
|  | Male | 45 (45.0) | 55 (55.0) |  |

### Service Availability and Readiness

Table 4 presents the bivariate associations between specific service availability and overall readiness. Five service domains were significantly associated with limited readiness. Absence of vaccination services showed the strongest crude association (OR = 16.05; 95% CI: 7.65–33.67; p<0.001): 87.0% of facilities without vaccination services had limited readiness, compared with 22.7% of those offering declared vaccination services. Absence of PMTCT services (OR = 9.83; 95% CI: 3.60–26.82; p<0.001), family planning (OR = 6.84; 95% CI: 2.21–21.16; p = 0.001), childbirth services (OR = 6.98; 95% CI: 3.01–16.22; p<0.001), and antenatal care (OR = 3.49; 95% CI: 1.06–11.55; p = 0.03) were each independently associated with limited readiness in bivariate analysis. Absence of surgery (p = 0.20) and blood transfusion (p = 0.42) services were not significantly associated with readiness.

**Table 4.**
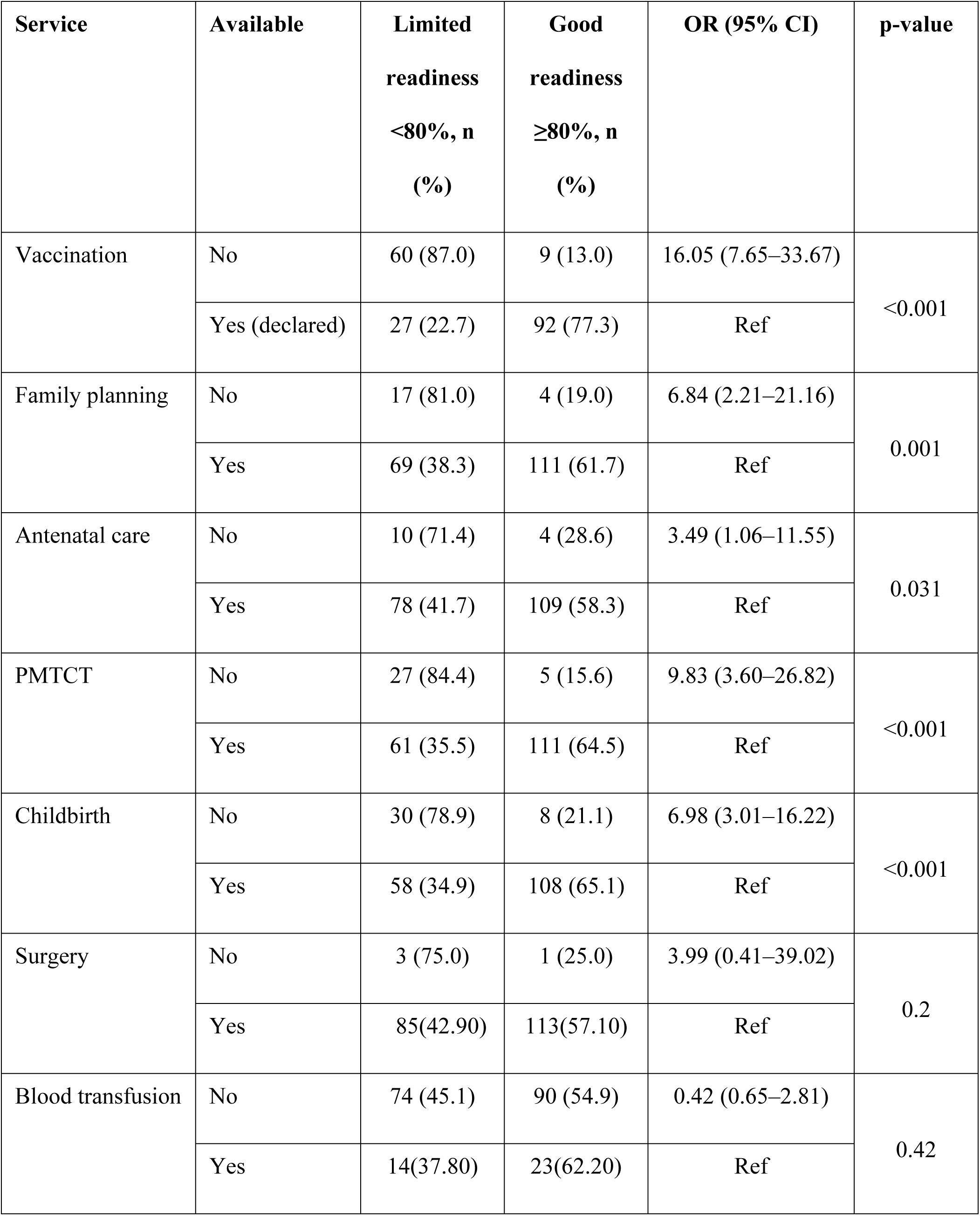
Association Between Service Availability and Good Readiness (≥80%)

**Table 5:**
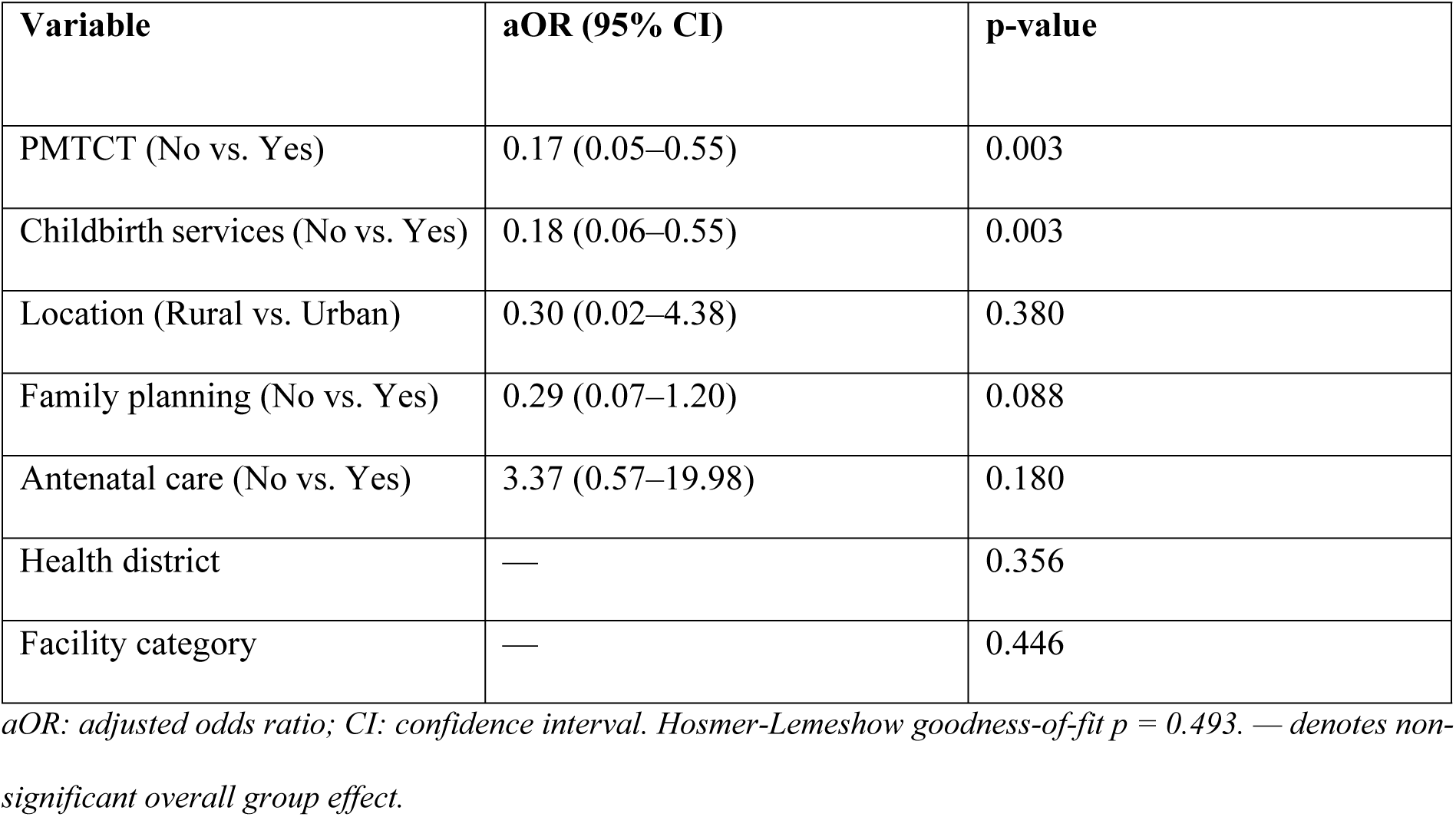
Multivariate Logistic Regression: Factors Independently Associated with Good Readiness (≥80%)

| Variable | aOR (95% CI) | p-value |
| --- | --- | --- |
| PMTCT (No vs. Yes) | 0.17 (0.05–0.55) | 0.003 |
| Childbirth services (No vs. Yes) | 0.18 (0.06–0.55) | 0.003 |
| Location (Rural vs. Urban) | 0.30 (0.02–4.38) | 0.380 |
| Family planning (No vs. Yes) | 0.29 (0.07–1.20) | 0.088 |
| Antenatal care (No vs. Yes) | 3.37 (0.57–19.98) | 0.180 |
| Health district | — | 0.356 |
| Facility category | — | 0.446 |

*aOR: adjusted odds ratio; CI: confidence interval. Hosmer-Lemeshow goodness-of-fit p = 0.493. — denotes non-* *significant overall group effect.*

### Factors Associated with Readiness: Multivariate Analysis

Using binary multiple logistic regression, only the absence of PMTCT and childbirth services remained independently associated with lower odds of good readiness. Facilities without PMTCT services had lower odds of achieving adequate readiness (aOR=0.17; 95% CI: 0.05–0.55; p=0.003), and those without childbirth services had lower odds (aOR=0.18; 95% CI: 0.06–0.55; p=0.003). Other variables, including district, facility category, and ownership, were no longer significant after adjustment, indicating that service availability explains much of the observed structural variation.

## Discussion

This study provides a comprehensive assessment of health facility readiness in Yaoundé, Cameroon, using the standardized SARA methodology. Our finding showed that 57.1% of health facilities met the 80% readiness threshold, indicating a moderate level of preparedness within the capital city. However, two factors; absence of PMTCT services and absence of childbirth/delivery services were independently associated with limited readiness, identifying maternal and child health service availability as a critical factor of broader health system functionality in this setting, highlighting significant gaps in health system capacity even within a major urban center.

### Overall Readiness in Context

The overall readiness score of 57.1% observed in Yaoundé is comparable to findings from other sub-Saharan African settings. In the Democratic Republic of the Congo, a SARA-based assessment of tuberculosis clinics in Lubumbashi found readiness levels of approximately 50% for providing diabetes diagnostic and treatment services[14]. Similarly, a nationally representative study in Ethiopia reported that general service readiness for non-communicable disease services varied considerably across regions and facility types, with significant gaps in lower-level facilities[15]. In Nepal, researchers found overall readiness scores of 59.9% across all health facilities, with tuberculosis services showing the lowest readiness (57.8%) and delivery and newborn care services the highest (67.1%)[16] . These comparisons suggest that the readiness levels observed in Yaoundé are consistent with broader patterns across low- and middle-income countries, where health systems struggle to maintain consistent operational capacity. Importantly, Côte d’Ivoire reported a higher readiness of 74.5% in its assessment of primary-level facilities for malaria management[17], underscoring that context-specific investments in particular service areas can meaningfully lift aggregate readiness

### The Central Role of Maternal and Child Health Services

The finding reveals a significant association between the absence absence of PMTCT services and absence of childbirth services and significantly lower overall facility readiness. Facilities without PMTCT services had lower odds of achieving the 80% readiness threshold (aOR=0.17; 95% CI: 0.05-0.55; p=0.003), while those without delivery services had 82% lower odds (aOR=0.18; 95% CI: 0.06-0.55; p=0.003).

This finding is consistent with evidence reported in Ethiopia, where Terefe and colleagues evaluated PMTCT service quality in public health facilities and found that overall performance was 74.09%, with input quality (including facility readiness) rated at only 60.4%[18]. Their study emphasized that availability of basic resources, adequate counseling, and reduced waiting times were critical determinants of service quality and client satisfaction. Similarly, in Mozambique, Dinis et al. demonstrated a significant association between service readiness and PMTCT cascade effectiveness, reinforcing that facilities with better readiness achieve superior prevention outcomes.

The strong association between delivery services and overall readiness observed in our study is consistent with findings from Nepal, where delivery and newborn care services demonstrated the highest readiness scores (67.1%) among all service domains[16]. This suggests that facilities investing in comprehensive obstetric care tend to have better overall operational capacity, possibly due to the intensive resource requirements and regulatory oversight associated with maternity services.

### Service Integration as a Marker of System Strength

Our findings are consistent with a broader hypothesis: PMTCT and delivery services act as *sentinel indicators* of systemic facility capacity rather than isolated service domains. Facilities that have established the infrastructure, staffing, consumables, and quality protocols required for these services appear also to have invested in the broader inputs that constitute readiness across all five SARA domains. This interpretation parallels findings from Ethiopia, where Tesema et al. demonstrated that readiness for communicable disease services was positively and independently associated with readiness for non-communicable disease services, indicating cross-cutting health system effects [15]. Their analysis revealed that increased readiness for communicable disease services was significantly associated with increased readiness for NCD services, suggesting that health system strengthening efforts have cross-cutting effects.

The integrated nature of health service delivery is further illustrated by innovative approaches in Cameroon itself. In conflict-affected regions, the Cameroon Baptist Convention Health Services has implemented the "Handshake Model," which uses vaccination as an entry point for delivering comprehensive care including malnutrition screening, antenatal care, deworming, and psychosocial support[19]. This model recognizes that families seek holistic care and that service integration increases community trust and uptake. While our study focused on urban Yaoundé rather than conflict zones, the principle that service integration reflects and reinforces system capacity remains relevant.

### Attenuation of district-level effects after adjustment

The bivariate analysis showed significant variation in readiness across health districts, with Efoulan (78.8%), Cité Verte (70.0%), and Nkolbisson (69.6%) performing substantially better than districts like Djoungolo (31.6%) and Nkoldongo (35.3%). However, these district-level differences lost statistical significance in the multivariate model once service availability (PMTCT and delivery) was accounted for. This finding suggests that the observed disparities between districts are largely explained by differential availability of these key maternal and child health services.

### Private Sector Dominance and Systemic Challenges

The dominance of private, secular facilities in our sample (90.2%) accurately reflects the healthcare landscape in Yaoundé. Interestingly, we found no significant difference in readiness between public and private facilities in the multivariate analysis. This suggests that in this urban context, the challenges to achieving readiness are systemic and affect all providers regardless of ownership.

This finding aligns with research from the Democratic Republic of the Congo, where Kakisingi and colleagues found that readiness varied significantly based on the managerial instance overseeing facilities[14]. However, unlike our study where ownership differences were not significant, their analysis revealed that public facilities and those with supplementary activity packages demonstrated different readiness profiles. The discrepancy may reflect Cameroon’s unique context where private secular facilities dominate and may have developed comparable operational systems to public facilities through market competition and regulatory requirements.

### Limitations

This study has several limitations. First, its cross-sectional design captures readiness at a single point in time and cannot establish causality. Second, readiness assessment measures potential capacity (inputs) rather than actual quality or outcomes of care. Third, self-reported data may introduce social desirability bias, although we attempted to mitigate this by combining interviews with direct observation of tracer items.

## Conclusions

This study reveals that nearly three-fifths of health facilities in Yaoundé meet the eighty percent readiness threshold, indicating moderate preparedness within the capital city, yet more than forty percent of facilities lack essential inputs for quality service delivery. The absence of Prevention of Mother-to-Child Transmission (PMTCT) services and childbirth/delivery services were independently associated with lower odds of good readiness; identifying these maternal and child health services as critical markers of broader health system functionality. Notably, district-level disparities in readiness were fully explained by differential availability of these services, suggesting that observed geographic variations reflect service gaps rather than inherent district characteristics. The lack of significant differences between public and private facilities further underscores that readiness challenges are systemic and require whole-system approaches.

These findings have direct policy implications for Cameroon’s progress toward universal health coverage. Investments in PMTCT and delivery services should be prioritized not only for their direct health benefits but also as potential catalysts for broader facility readiness improvements.

Regulatory and quality improvement mechanisms must engage the dominant private sector, and the eighty percent threshold provides an evidence-based benchmark for monitoring national health system strengthening. Addressing systemic structural imbalances illustrated by the Efoulan Health District serving twenty times its recommended population requires sustained, multi-sectoral investment. Strengthening core inputs required for maternal and child health services may serve as an effective entry point for broader facility readiness improvements. Cameroon can make substantial progress toward equitable, quality healthcare for all citizens.

## Data Availability

The following existing data sources were used: "Nguefack-Tsague and Tedjo_Final.xlsx" from Data Mendeley available via 10.17632/myfzt2tkvf.1.

https://doi.org/10.17632/myfzt2tkvf.1

## Acknowledgements

The authors would like to thank all health facility managers and healthcare professionals who participated in this study and generously provided their time and information. We also acknowledge the support of the district health authorities and the Ministry of Public Health of Cameroon for facilitating data collection

